# Genomic epidemiology of SARS-CoV-2 in Colombia

**DOI:** 10.1101/2020.06.26.20135715

**Authors:** Katherine Laiton-Donato, Christian Julián Villabona-Arenas, José A. Usme-Ciro, Carlos Franco-Muñoz, Diego A. Álvarez-Díaz, Liz Stephany Villabona-Arenas, Susy Echeverría-Londoño, Zulma M. Cucunubá, Nicolás D. Franco-Sierra, Astrid C. Flórez, Carolina Ferro, Nadim J. Ajami, Diana Marcela Walteros, Franklin Prieto, Carlos Andrés Durán, Martha Lucia Ospina-Martínez, Marcela Mercado-Reyes

## Abstract

Coronavirus disease 2019 (COVID-19) was first diagnosed in Colombia from a traveler arriving from Italy on February 26, 2020. To date, available data on the origins and number or introductions of SARS-CoV-2 into the country are limited. Here, we sequenced SARS-CoV-2 from 43 clinical samples and—together with other 73 genomes sequences available from the country—we investigated the emergence and the routes of importation of COVID-19 into Colombia using epidemiological, historical air travel and phylogenetic observations. Our study provided evidence of multiple introductions, mostly from Europe, with at least 12 lineages being documented. Phylogenetic findings validated the lineage diversity, supported multiple importation events and the evolutionary relationship of epidemiologically-linked transmission chains. Our results reconstruct the early evolutionary history of SARS-CoV-2 in Colombia and highlight the advantages of genome sequencing to complement COVID-19 outbreak investigation.

## Introduction

Coronavirus disease 2019 (COVID-19) is a life-threatening respiratory illness caused by SARS-CoV-2, an emerging zoonotic virus first identified in Wuhan city, Hubei province, China (1). The first confirmed cases of COVID-19 were reported on January 12, 2020, from patients presenting respiratory symptoms between December 8, 2019 and January 2, 2020 (2). Despite early containment and mitigation measures set in place (3), the high infectiousness, presymptomatic and prolonged transmission of SARS-CoV-2 (4,5) combined with other factors such as globalization, led to its rapid spread across the world.

Rigorous contact-tracing and physical distancing measures implemented in different countries have been demonstrated to be effective in delaying the epidemic during the contention phase (6–9). However, ensuing lockdowns and travel restrictions to minimize the healthcare burden has led to wellbeing decline and economic downturn, with profound impacts in low-to-middle income countries (10). The contention phase in Colombia started with the first case of COVID-19 confirmed by the Instituto Nacional de Salud (INS) on March 6, 2020, from a person returning from Italy on February 26, 2020 (11). On March 23, 2020, 314 cases had been confirmed which prompted the closure of all the country borders to contain the outbreak. “On March 31, 2020, more than 10% of confirmed cases were individuals with no known exposure to a patient with COVID-19, presumably due to extensive community transmission, and therefore the mitigation phase was declared with physical-distancing as the main strategy to further limit virus spread.” On June 18, 2020, 57,046 confirmed cases, and 1,864 deaths had been reported in Colombia (12).

The unprecedented global health and societal emergency posed by the COVID-19 pandemic urged data sharing and faster-than-ever outbreak research developments which are reflected in the fact that over 37.000 SARS-CoV-2 complete genomes have been made available through public databases, mainly GISAID (Global Initiative on Sharing All Influenza Data). SARS-CoV-2 is an RNA virus with an estimated substitution rate of around 0.8×10−3 to 1.1×10−3 (13,14) which means that it evolves fast as it is transmitted. The availability of SARS-CoV-2 genomes enable us to detect this fast generating variation and therefore genomic epidemiology is a powerful approach to characterize the outbreak (15). Genomic epidemiology which on phylogenetic analysis and has allowed researchers across the world to ascertain SARS-CoV-2 emergence in humans, to reveal the importation and local transmission chains not detected by travel history and traditional contact-tracing strategies and, to trace the geographic spread and prevalence of strains bearing specific mutations of epidemiologic relevance (16–20).

Here we describe the complete genome sequencing of SARS-CoV-2 from 43 clinical samples together with the epidemiological investigation of imported cases and the phylogenetic findings using 122 genome sequences from Colombia, which characterize the epidemic onset of COVID-19 in the country. Our study suggests multiple independent introductions from across the world, mainly from Europe and the co-circulation of multiple lineages in the most affected departments. These findings contribute to the molecular surveillance of COVID-19 and provide venues for future outbreak response and interventions in the country.

## Materials and methods

### Sample preparation

Nasopharyngeal swabs samples from patients with clinical presentations of SARS-CoV-2 across the country were received at the Instituto Nacional de Salud (INS, National Institute of Health) as part of the virological surveillance of COVID-19. Diagnosing suspected cases of COVID-19 was performed by using the qRT-PCR protocol (21) recommended and transferred by the PAHO/WHO to the INS. Due to scarce resources, a total of 43 samples (Suppl Table 1) were selected for genome sequencing representing either the earliest documented samples per affected department (at least one sample from each department) or samples linked to transmission chains. Viral RNA extraction was performed by using the QIAamp Viral RNA Mini kit (Qiagen Inc., Chatsworth, CA, USA) or the MagNA Pure LC nucleic acid extraction system (Roche Diagnostics GmbH, Mannheim, Germany).

### Genomic library preparation and sequencing

Library preparation and sequencing were performed following the ARTIC network (real-time molecular epidemiology for outbreak response) protocol and using both Nanopore and Illumina technologies (22). Ten samples were processed using Oxford Nanopore Technologies (Oxford Nanopore Technologies, Oxford, UK). The remaining 33 samples were processed with the Nextera XT DNA library prep kit (Illumina, San Diego, CA, USA) and sequenced using the MiSeq reagent kit version 2 and the MiSeq instrument (Illumina, San Diego, CA, USA).

### Genomic sequence assembly

Nanopore reads were basecalled using Guppy version 3.2.2 (Oxford Nanopore Technologies, Oxford, UK) and then demultiplexed and trimmed using Porechop version 0.3.2_pre (23). Processed reads were aligned against SARS-CoV-2 reference genome (GenBank NC_045512.2) using BWA-MEM (24). Single nucleotide variants were called with a depth of at least 200x and then polished consensus was generated using Nanopolish version 0.13.2 (25). MiSeq reads were demultiplexed and quality control was performed with a Q-score threshold of 30 using fastp (26). Processed reads were aligned against SARS-CoV-2 reference genome (GenBank NC_045512.2), single nucleotide variants were called with a depth of at least 100x and consensus genomes were generated using BWA-MEM version 0.7.17 (24) and BBMap (27).

### Phylogenetic analysis of SARS-CoV-2 in Colombia

The SARS-CoV-2 genome sequences obtained in the present study and those from Colombia deposited in GISAID were collated (n=122) and combined with 1461 representative genome sequences from the South America-focused subsampling available from NextStrain (28) as of 20th May 2020 (Table S2) plus reference MN908947.3. The median value of sequences available per department was 1.5 (mean of 3.9 sequences, a max of 45 sequences were obtained from a single locality). The full genomic dataset was classified in lineages (29) using PANGOLIN (Phylogenetic Assignment of Named Global Outbreak LINeages) (30) and aligned with 10 iterative refinements using MAFFT (31). All the alignment positions flagged as problematic for phylogenetic inference were removed (e.g. highly homoplasic positions and 3’ and 5’ ends) (32). Maximum likelihood phylogenetic reconstruction was performed with the curated alignment and a HKY+Γ4 substitution model (33) using IQTREE (34). Branch support was estimated with an SH-like approximate likelihood ratio test (SH-aLRT)—a value greater than or equal to 0.75 was considered a high SH-aLRT (35). Six sequences from Colombia that showed inconsistent temporal signal with a clock analysis using TreeTime (36) were removed from further analysis. Time-scaled trees were inferred and rooted with least-squares criteria and the evolutionary rate of about 1.1*10^-3^ subs/site/year estimated by (13) using TreeTime (36) and LSD (37). The geographical locations (aggregated by continent except for Colombia) of the sequence data were considered as discrete states and used for migration inference—with transitions modeled as a time reversible process—in TreeTime (36). The number of state transitions into Colombia were interpreted as a proxy for the minimum number of introductions. In sensitivity analysis and in order to measure the effect of the SARS-CoV-2 uneven genomic representativeness across the world, two down sampling strategies datasets were implemented where, based on location, the sequences were randomly resampled 100 times and the phylogenetic and migration inference was replicated. The downsampling strategies were as follows: (i) retaining a number of sequences per region (whenever possible) equal to the number of sequences available for Colombia and (ii) retaining 50 sequences per region and the total number of sequences from Colombia (this was the most even sampling per region for the South America-focused subsample).

### Potential routes of COVID-19 importation in Colombia

The relative proportion of expected importations of COVID-19 by country to Colombia was inferred by taking into account SARS-CoV-2 incidence per international air passengers arriving in Colombia and the available flight travel. The number of international flights and number of passengers arriving to Colombia from January 1 to March 9, 2020 were obtained from the Special Administrative Unit of Civil Aeronautics of Colombia (Aerocivil, aerocivil.gov.co). The air travel data consists of direct flights from 14 countries to 7 main cities. COVID-19 incidence for each of the 14 countries with direct flights to Colombia was calculated using the number of confirmed cases reported by the World Health Organization—as of March 17, 2020, the date when travel restrictions started in Colombia (38)—and the total population for each country for 2019 reported in the United Nations World Population Prospects 2019 database (39) as in (40) (Appendix).

## Results and discussion

### Epidemiology investigation of introductions, contact-tracing and community transmission of SARS-CoV-2 in Colombia

In Colombia, preventive isolation and monitoring for passengers arriving from China, Italy, France, and Spain started on March 10, 2020. National health emergency was declared on March 12 and tougher measures started to be set in place including the closing of borders (March 17) the ban of international (March 20) and domestic flights (March 25). Implementations of lockdowns occurred from March 25 onwards (Resolutions 380 and 385 from the Colombian Ministry of Health and Social Protection; Decree 412 and 457 from the Colombian Ministry of the Interior; Decree 439 from the Ministry of Transport). Despite massive drop in air traffic, more than 15.500 residents returned to Colombia through humanitarian flights during April and June (41).

As per June the first, over 30 thousand cases of COVID-19 had been documented in the country and 857 cases (2.8%) had been linked to travel abroad (Figure 1A). The majority of imported cases were symptomatic (n=816, 95.2%) and the most important geographical sources were Spain (n=245, 28.6%), the United States of America (n=203, 23.7%), Ecuador (n=50, 5.8%), Mexico (n=49, 5.7%) and Brazil (n=41, 4.8%). The 41 asymptomatic imported cases were detected through contact-tracing and the majority of these cases were imported from Spain (n=16, 39%), the United States (n=13, 31.7%), Brazil (n=3, 7.3%) and Mexico (n=2, 4.9%). Most of the symptomatic imported cases were traced back to European countries and then to countries in the Americas. The most important geographical sources in terms of imported cases were Spain (30.5%), the United States of America (25.2%), Mexico (6%), Ecuador (5.8%), and Brazil (5.1%). The number of symptomatic imported cases steadily increased and reached a peak on March 14, 2020, when local cases were on the rise and earlier than the closure of borders and ban of international air travel. Our estimate is based on the average incubation time of COVID-19 (42) and consequently 4.8 days earlier than peak based on symptoms onset (March 18, Figure 1B). The first introductions were predominantly linked to Europe, however, both Europe and the American region were important geographical sources of infections during the onset of the epidemic. The introductions post-peak occurred for the most part from South American countries.

**Figure 1.**
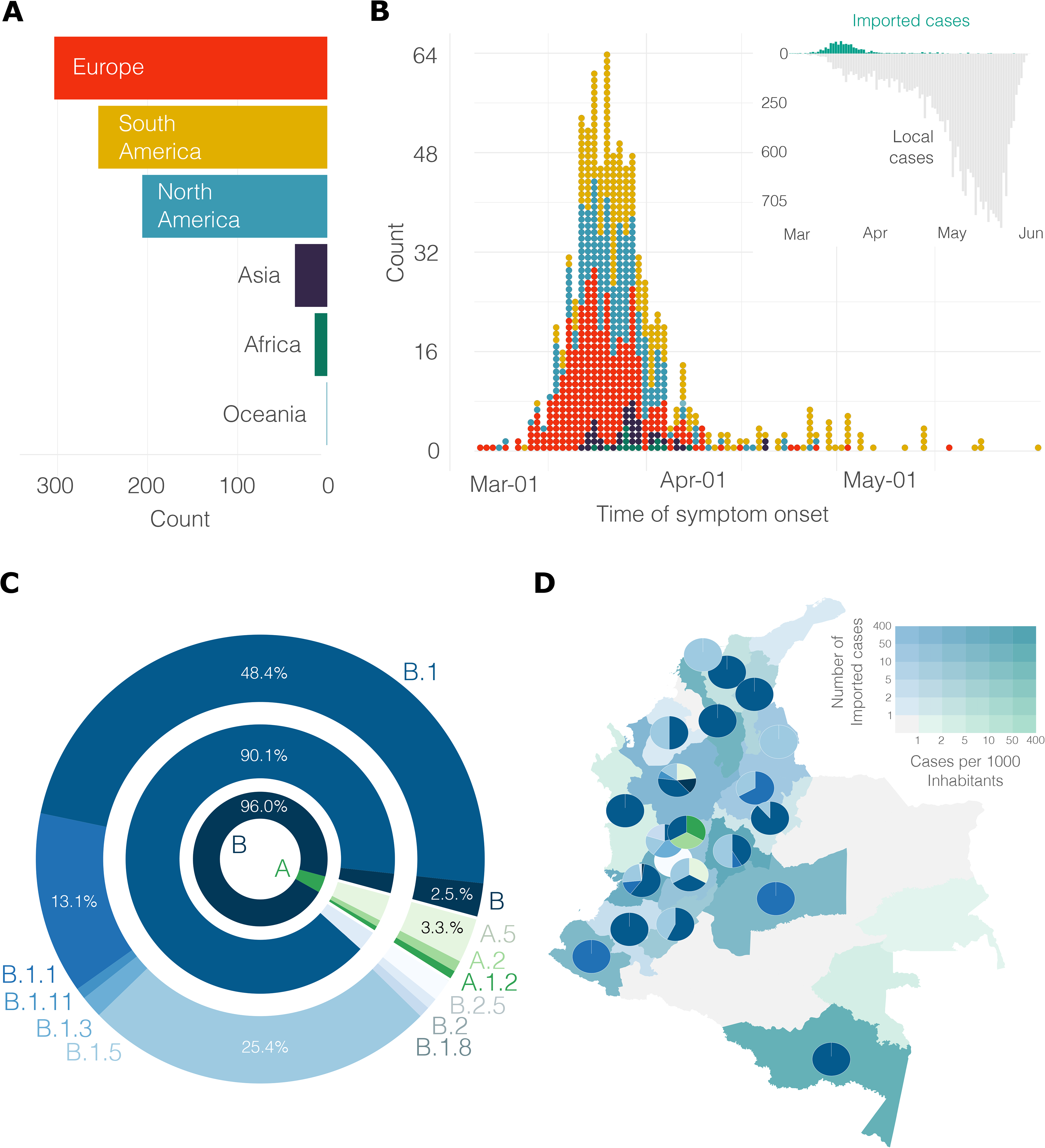
Proportion of imported and local cases early during the pandemic of COVID-19 in Colombia. A) Region of origin for the reported imported cases. B) Distribution over time of the symptomatic imported and local cases colored by region (as in A).

### SARS-CoV-2 diversity in Colombia

In order to gain a better understanding of the dynamics of SARS-CoV-2 spread into Colombia, 43 whole genome sequences obtained in the present study were combined with sequences from Colombia deposited in GISAID, resulting in a set of 122 complete genomes. Sequences from Colombia were classified in a total of 12 sublineages: A.1.2, A.2, A.5, B, B.1, B.1.1, B.1.3, B.1.5, B.1.8, B. 1.11, B.2, and B.2.5. The proportion of lineages documented in Colombia seems to reflect founder effects, for example, sublineages B.1, B.1.1, and B.1.5 were found in the early epidemiologically-linked transmission chains and, consistently, the most frequently observed lineages in the genome sequence data from Colombia were B.1 (n= 59, 48.4%), B.1.5 (n=31, 25.4%) and B.1.1 (n=16, 13.1%) (Figure 2A). Comparable findings were observed for other South American countries (43–45) from the South America-focused subsampling available from NextStrain, where the most frequently observed lineages were B.1 (n= 149, 60.8%), B.1.5 (n=33, 13.5%) and A.5 (n=14, 5.7%). Colombia is divided into 32 departments and the genome sequence data covered the 20 affected departments (up to date) and the capital district. The median value of available sequences per department was 1.5 (mean of 3.9 sequences, a max of 45 sequences were obtained from a single locality). Consequently, on average, one single lineage was identified by department. For instance, the number of documented lineages was highly correlated with the availability of samples (Pearson’s product-moment correlation of 0.72, *p*-value<0.001) and uncorrelated with the number of local cases (Pearson’s product-moment correlation of 0.35, *p*-value=0.049). Five different lineages were documented in the departments of Valle del Cauca and Antioquia and three different lineages were documented in Cundinamarca which are the departments with a higher number of samples (Figure 2B) and whose capitals are the most populated. A moderate positive correlation was observed between the number of lineages documented in a department and the number of imported cases (Pearson’s product-moment correlation 0.51, *p*-value = 0.002).

**Figure 2.**
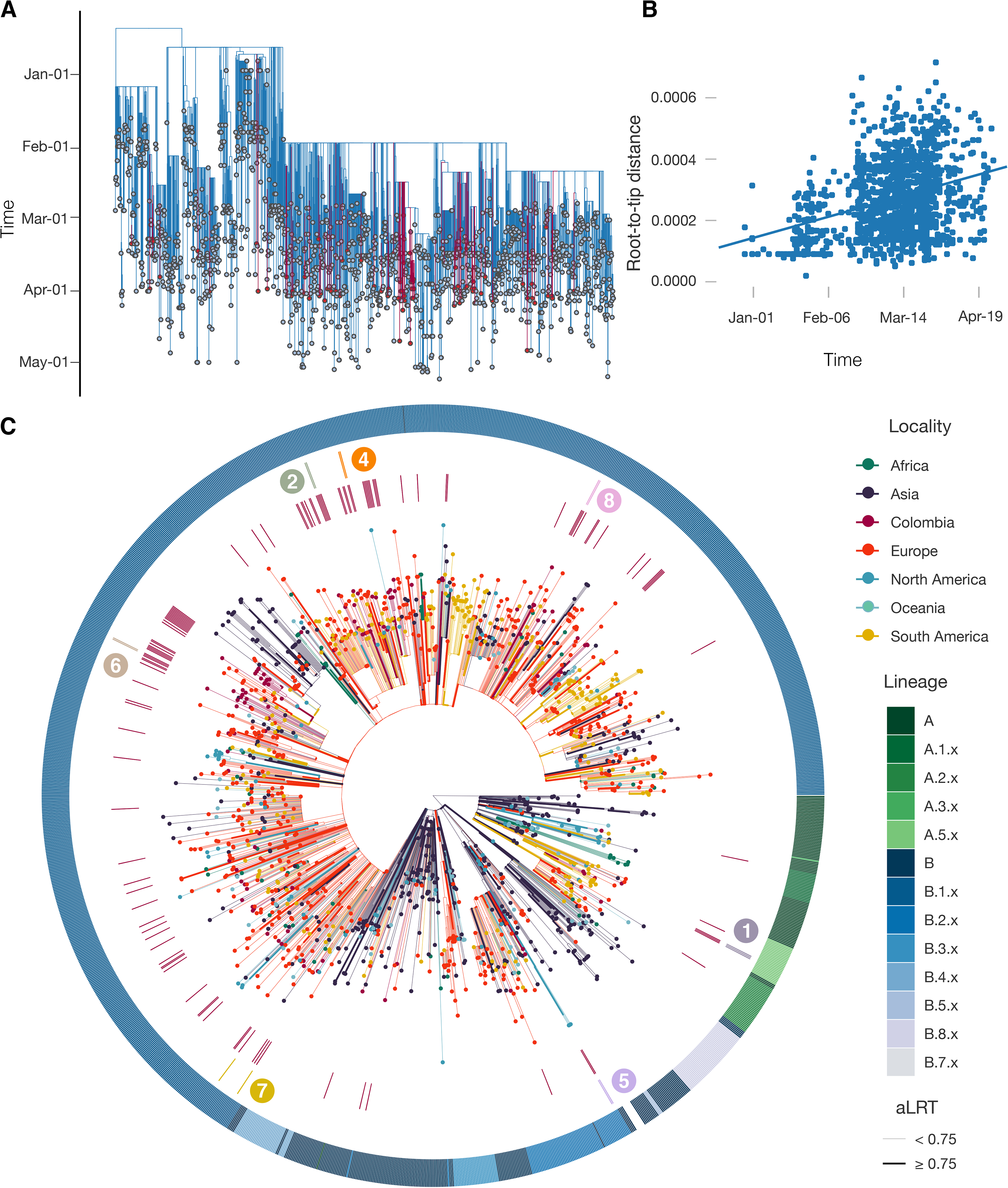
Frequency and distribution of SARS-CoV-2 lineages in Colombia. A) Frequency of the lineages and sub-lineages of SARS-CoV-2 identified in Colombia. B) Distribution of lineages across the country; departments are colored by the number of imported cases per 10 thousand inhabitants and the number of reported introductions.

### Molecular evolution of SARS-CoV-2 in Colombia

A total of 133 nucleotide variants (NVs) were identified using the full genome sequences from Colombia and the reference NC_045512.2. The majority (n=131) of the NVs fell in the coding region (one NV was identified at each non-coding end). A total of 71 (54,2%) of the NVs in coding sites led to non-synonymous substitutions. Most of the NVs (92 out of 133) were unique to a sequence and among the shared NVs, 38 (out of 41) were associated with a specific lineage (Tables S3 and S4). These observations suggest that the substitutions are not laboratory-specific and most likely the outcome of *in-situ* evolution and/or shared ancestry (Appendix).

In our study, 90% sequences from Colombia displayed amino acid change D614G (108 of a total 120 Colombian sequences covered the region), while the remaining 10% (12 sequences) displayed the D614 (Table S4). G614 has been associated with higher infectivity (46) and greater transmissibility (47) with no effects on disease severity outcomes (48). All G614 sequences also carried mutations that segregate together as described in (47): at the 5’UTR we identified the nucleotide substitution C241T, at the ORF1ab (Nsp3 encoding-gene) the synonymous substitution C3037T and at the ORF1ab (RdRp encoding-gene) the amino acid change P4715L position. The presence of these and other mutations warrant further monitoring because they can be phenotypically and epidemiologically relevant.

Most of the patients (n=90) from Colombia for whom genomic sequences were available were symptomatic (59.6% with cough and fever and the remaining with at least one of these two symptoms) and (n=10) of them died (70% presented comorbidities) (Table S1). However, given the limited number of available genomic sequences, it was not possible to reliably investigate any genetic determinant of clinical outcome.

### Evolutionary relationships between local and global SARS-CoV-2 isolates

The time-stamped phylogeny of 122 isolates of Colombia and 1462 representative global SARS-CoV-2 isolates (Figure 3A) showed that the estimated time to the most recent common ancestor for the sampled sequence data is December 7, 2019 (October 25 - December 26). Asia was the inferred ancestral state at the root. Both these observations are in line with the known epidemiology of the pandemic. A root-to-tip regression of genetic distance against sampling time evidenced consistent temporal signal in the sequence data (Figure 3B). The isolates from Colombia appeared intersperse among the isolates from other countries (Figure 3A and 3C) suggesting multiple introductions. However, there was considerable phylogenetic uncertainty along the tree and the fine-grained relationships of the isolates from Colombia could not be resolved with confidence (Figure S1). Phylogenetic uncertainty together with uneven sampling made the quantification of the number of introductions into the country challenging, let alone dating the time of the introductions. The number of state transitions into Colombia (Figure 4A) heavily relies on the number and nature of the sequences that are included from other locations. Using all sequences in the South America-focused subsampling available from NextStrain, we estimated that on average 64 (IQ 62-67) introductions have occurred into the country but this estimate gets lower as we reduce the number of samples (sensitivity analyses) from other locations (down to 22 with the most even down-sampled dataset). Independently of the dataset (either the complete or the subsampled datasets) and in line with the epidemiological information, most of the geographical source attributions of the introductions are attributed to Europe (Figure 4B and Figure S2). This observation is also in agreement with our estimates using travel data as detailed below (Figure 4C and Figure S2).

**Figure 3.**
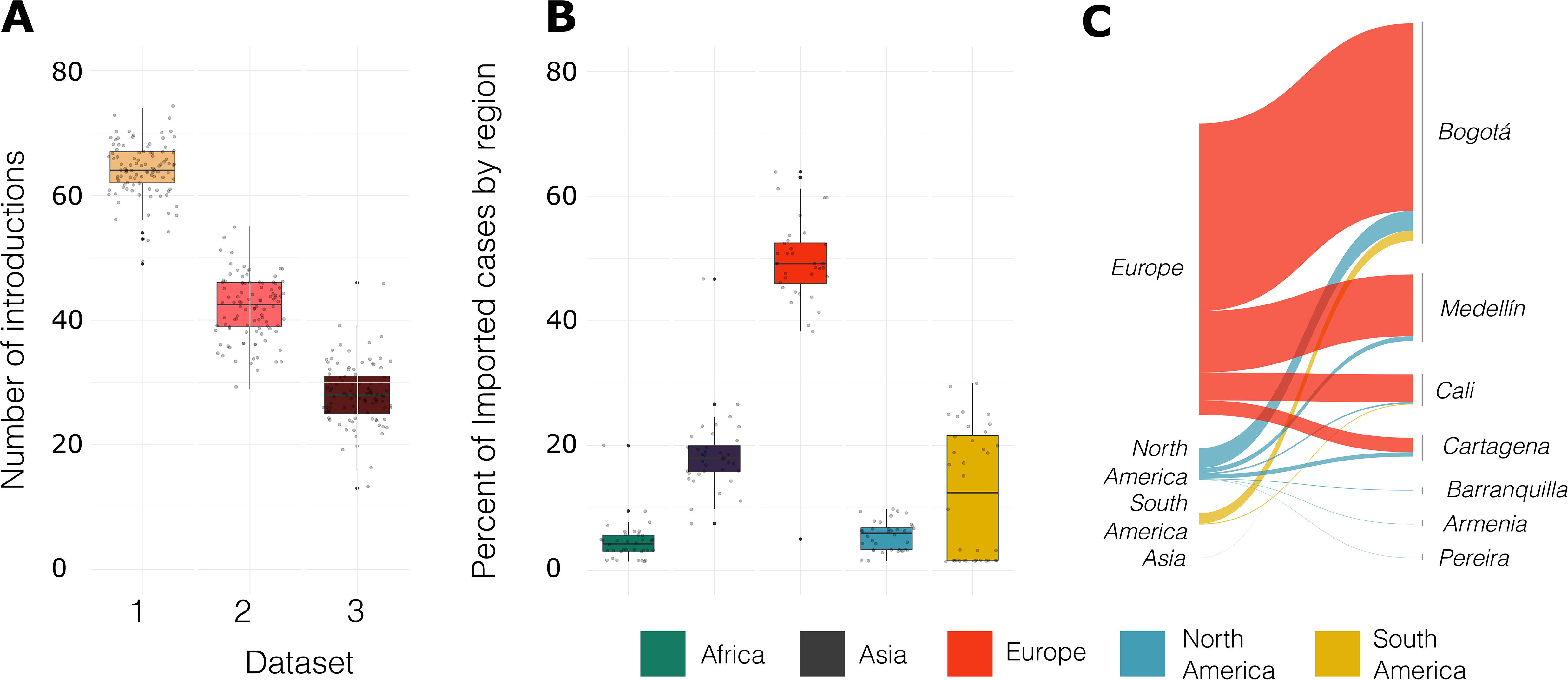
Phylogeny of SARS-CoV-2 from Colombia. A) Time-resolved maximum likelihood tree of 1578 SARS-CoV-2 sequences including 122 from Colombia (red). B) Root-to-tip distance regression for the sequence data in A. C) Time-resolved maximum likelihood annotated by region of isolation. The outer ring represents SARS-CoV-2 lineages. The inner red ring highlights the relative position of the sequences from Colombia. The middle ring and the corresponding numbers indicate sequences from epidemiologically-linked transmission chains. Branches are colored by the geographical attribution from the migration inference. Highly supported groups are delineated by thicker solid lines. A detailed maximum likelihood tree is available at itol.embl.de/tree/8619015795401231596483440.

Between January and March 2020, 7 cities of Colombia received 1,593,211 international passengers from 14 countries. Bogotá concentrated the majority of the flights with around 77% of the passengers, followed by Medellín with 11%, Cartagena with 6%, and Cali with 4%. In total, 35% of international passengers started their journey in the USA, 17% in Mexico, and 12% in Chile. However, we estimate that 87% of all imported COVID-19 cases came from Europe, 9.5% from North America, and 3.4% from South America. When stratified by country, the primary source of importation was Spain with 71.4% (Figure S2).of the cases, followed by the USA with 8.4%, Germany with 8%, and France with 3.4% 65.2% of the COVID-19 cases were expected to arrive at Bogotá, followed by Medellín with 20% and Cali with 9%. We estimate that the route Spain-Bogotá carried 42% of the total imported cases.

Since the first introduction of SARS-CoV-2 in Colombia (February 26, 2020) contact-tracing efforts had been set in place. Multiple sequences were obtained from seven distinct early epidemiologically-linked transmission chains (Table S1). This information was mapped into the phylogeny (Figure 3C) and all but one set of sequences did not group. However, it appeared very close in the tree. This underscores the potential utility of genomic epidemiology to link individuals with incomplete information (e.g. cases that end-up disconnected due to intermediate asymptomatic carriers) and complement outbreak transmission.

This study has some limitations. Firstly, the geographical sources of infection relied on people to self-report their own symptoms onset and travel history, which is subject to inaccuracies. Secondly, we use air travel data from important destinations in Colombia but other locations may also have fueled COVID-19 emergence and dissemination in the country; flight travel data was not available from March 9, 2020, onwards. Thirdly, the number of sequences sampled represented a tiny fraction of the documented number of imported cases into Colombia; the sample was selected to be a country-wide representative given limited resources for genome sequencing and consequently, the introduced viral diversity may also have been underestimated. Another limitation of this analysis is the inherent uncertainty stemming from global unsystematic sampling. Therefore, the inferences about the number of introductions and the corresponding geographical source need to be interpreted with caution. We attempted to overcome this by undertaking sensitivity analyses and contrasting the results with the available epidemiological data and our estimates from travel data. However, more sequence data from Colombia and undersampled countries together with information of sampling representativeness per country is needed in order to account for sampling uncertainty in a more statistically rigorous manner.

Our study provided evidence that an important number of independent introductions occurred to Colombia with at least 12 lineages documented. Most of the notified introductions to Latin American countries occurred from Europe and this observation was supported by phylogenetic and air travel data (49,50). Although the sequence data do not represent the actual number of epidemiologically-linked transmission chains, our phylogenetic findings validated the linkage for those epidemiologically-linked transmission chains with available sequence data. Our results further underscore the advantages of genome sequencing to complement COVID-19 outbreak investigation and support the need for a more comprehensive country-wide study of the epidemiology and spread of SARS-CoV-2 in Colombia.

## Data Availability

Data supporting this paper can be accessed through GISAID at https://www.gisaid.org/ here
and used under a Creative Commons Attribution (CC-BY) licence

## Acknowledgments

The authors thank the National Laboratory Network and Virology Group of INS for routine virologic surveillance of SARS-CoV-2 in Colombia. The authors thank all researchers who deposited genomes in GISAID’s EpiCoV (TM) Database contributing to genomic diversity and phylogenetic relationship of SARS-CoV-2. The authors thank Maylin Gonzalez Herrera for her technical assistance.

## Ethical statement

According to the national law 9/1979, decrees 786/1990 and 2323/2006, the Instituto Nacional de Salud is the reference lab and health authority of the national network of laboratories and in cases of public health emergency or those in which scientific research for public health purposes as required, the Instituto Nacional de Salud may use the biological material for research purposes, without informed consent, which includes the anonymous disclosure of results. The information used for this study comes from secondary sources of data that were previously anonymized and do not represent a risk to the community.

## Author Bio

Katherine Laiton-Donato is the head of the Sequencing and Genomics Unit, Dirección de Investigación en Salud Pública, Instituto Nacional de Salud, Colombia. Her research is focused on molecular virology of emerging viruses with impact in public health.

---

**Figure 4. Potential routes of COVID-19 importation in Colombia**. A) The number of transition changes into Colombia following migration inference using all available sequences per region [dataset 1], retaining a number of sequences per region (whenever possible) equal to the number of sequences available for Colombia [dataset 2] and 50 sequences per region and all sequences from Colombia [dataset 3]. B) Geographical source attribution for every transition into Colombia derived from the migration inference using all the available sequences per region. C) Geographical contribution inferred using Air travel data per country aggregated by region.

**Table S1**. Metadata of COVID-19 diagnosed patients in Colombia.

**Table S2**. GISAID’s nCoV-19 Acknowledgements.

**Table S3**. Nucleotide substitution patterns of the different lineages of SARS-CoV-2 circulating in Colombia.

**Table S4**. Amino acid change patterns of the different lineages of SARS-CoV-2 circulating in Colombia.

**Table S5**. Estimates of evolutionary divergence of SARS-CoV-2 over sequence pairs within and between lineages/sublineages from Colombia.

**Figure S1. Example of a clade with sequences from a local transmission chain from Colombia**. Of note, fine-grained relationships could not always be resolved with confidence. Branch supports (approximate likelihood ratio test, aLRT) below 0.75 are not shown. Tips are labeled by country (and department in the case of isolates from Colombia) and colored by region. The scales are proportional to the number of substitutions per site.

**Figure S2. Potential countries of introductions of SARS-CoV-2**. A) Geographical source attribution for every transition into Colombia derived from the migration inference retaining a number of sequences per region (whenever possible) [dataset 2] equal to the number of sequences available for Colombia and retaining 50 sequences per region and all sequences from Colombia [dataset 3]. B) Number of passengers arriving in Colombia from different countries. C) Geographical contribution inferred using Air travel data per country.

## References

1. Wu F, Zhao S, Yu B, Chen Y-M, Wang W, Song Z-G, et al. A new coronavirus associated with human respiratory disease in China [Internet]. Vol. 579, Nature. 2020. p. 265–9. Available from: http://dx.doi.org/10.1038/s41586-020-2008-3

2. WHO. Novel Coronavirus – China [Internet]. 2020 [cited 2020 Jun 16]. Available from: https://www.who.int/csr/don/12-january-2020-novel-coronavirus-china/en/

3. Kraemer MUG, Yang C-H, Gutierrez B, Wu C-H, Klein B, Pigott DM, et al. The effect of human mobility and control measures on the COVID-19 epidemic in China. Science. 2020 May 1;368(6490):493–7.

4. He X, Lau EHY, Wu P, Deng X, Wang J, Hao X, et al. Temporal dynamics in viral shedding and transmissibility of COVID-19. Nat Med. 2020 May;26(5):672–5.

5. Li J, Zhang L, Liu B, Song D. Case Report: Viral Shedding for 60 Days in a Woman with COVID-19. Am J Trop Med Hyg. 2020 Jun;102(6):1210–3.

6. Jarvis CI, Van Zandvoort K, Gimma A, Prem K, CMMID COVID-19 working group, Klepac P, et al. Quantifying the impact of physical distance measures on the transmission of COVID-19 in the UK. BMC Med. 2020 May 7;18(1):124.

7. Prem K, Liu Y, Russell TW, Kucharski AJ, Eggo RM, Davies N, et al. The effect of control strategies to reduce social mixing on outcomes of the COVID-19 epidemic in Wuhan, China: a modelling study [Internet]. Vol. 5, The Lancet Public Health. 2020. p. e261–70. Available from: http://dx.doi.org/10.1016/s2468-2667(20)30073-6

8. Hellewell J, Abbott S, Gimma A, Bosse NI, Jarvis CI, Russell TW, et al. Feasibility of controlling COVID-19 outbreaks by isolation of cases and contacts. Lancet Glob Health. 2020 Apr;8(4):e488–96.

9. Chinazzi M, Davis JT, Ajelli M, Gioannini C, Litvinova M, Merler S, et al. The effect of travel restrictions on the spread of the 2019 novel coronavirus (COVID-19) outbreak. Science. 2020 Apr 24;368(6489):395–400.

10. Ribeiro F, Leist A. Who is going to pay the price of Covid-19? Reflections about an unequal Brazil [Internet]. Vol. 19, International Journal for Equity in Health. 2020. Available from: http://dx.doi.org/10.1186/s12939-020-01207-2

11. MINSALUD. Minsalud confirma seis nuevos casos de coronavirus (COVID-19) en Colombia [Internet]. Boletín de Prensa No 057 de 2020. Bogotá, Colombia; 2020 [cited 2020 May 24]. Available from: https://www.minsalud.gov.co/Paginas/Minsalud-confirma-seis-nuevos-casos-de-coronavirus-(COVID-19)-en-Colombia.aspx

12. INS. Coronavirus (COVID - 2019) en Colombia [Internet]. Instituto Nacional de Salud; 2020 [cited 2020 Jun 2]. Available from: https://www.ins.gov.co/Noticias/Paginas/Coronavirus.aspx

13. Duchene S, Featherstone L, Haritopoulou-Sinanidou M, Rambaut A, Lemey P, Baele G. Temporal signal and the phylodynamic threshold of SARS-CoV-2 [Internet]. Available from: http://dx.doi.org/10.1101/2020.05.04.077735

14. Worobey M, Pekar J, Larsen BB, Nelson MI, Hill V, Joy JB, et al. The emergence of SARS-CoV-2 in Europe and the US. bioRxiv [Internet]. 2020 May 23; Available from: http://dx.doi.org/10.1101/2020.05.21.109322

15. Grubaugh ND, Ladner JT, Lemey P, Pybus OG, Rambaut A, Holmes EC, et al. Tracking virus outbreaks in the twenty-first century. Nat Microbiol. 2019 Jan;4(1):10–9.

16. Dellicour S, Durkin K, Hong SL, Vanmechelen B, Martí-Carreras J, Gill MS, et al. A phylodynamic workflow to rapidly gain insights into the dispersal history and dynamics of SARS-CoV-2 lineages [Internet]. Available from: http://dx.doi.org/10.1101/2020.05.05.078758

17. Fauver JR, Petrone ME, Hodcroft EB, Shioda K, Ehrlich HY, Watts AG, et al. Coast-to-coast spread of SARS-CoV-2 in the United States revealed by genomic epidemiology. medRxiv [Internet]. 2020 Mar 26; Available from: http://dx.doi.org/10.1101/2020.03.25.20043828

18. Lu J, du Plessis L, Liu Z, Hill V, Kang M, Lin H, et al. Genomic Epidemiology of SARS-CoV-2 in Guangdong Province, China. Cell. 2020 May 28;181(5):997–1003.e9.

19. Eden J-S, Rockett R, Carter I, Rahman H, de Ligt J, Hadfield J, et al. An emergent clade of SARS-CoV-2 linked to returned travellers from Iran. Virus Evol. 2020 Jan;6(1):veaa027.

20. Gudbjartsson DF, Helgason A, Jonsson H, Magnusson OT, Melsted P, Norddahl GL, et al. Spread of SARS-CoV-2 in the Icelandic Population. N Engl J Med. 2020 Jun 11;382(24):2302–15.

21. Corman VM, Landt O, Kaiser M, Molenkamp R, Meijer A, Chu DK, et al. Detection of 2019 novel coronavirus (2019-nCoV) by real-time RT-PCR. Euro Surveill [Internet]. 2020 Jan;25(3). Available from: http://dx.doi.org/10.2807/1560-7917.ES.2020.25.3.2000045

22. Quick J. nCoV-2019 sequencing protocol [Internet]. protocols.io; 2020a [cited 2020 Mar 2]. Available from: https://www.protocols.io/view/ncov-2019-sequencing-protocol-bbmuik6w

23. Wick R. rrwick/Porechop [Internet]. 2020 [cited 2020 Jun 18]. Available from: https://github.com/rrwick/Porechop

24. Li H, Wong CK, Ge N, Mori Y. bwa - Burrows-Wheeler Alignment Tool [Internet]. 2020. Available from: http://bio-bwa.sourceforge.net/bwa.shtml

25. Simpson J. Nanopolish: Signal-level algorithms for MinION data. Github Available at: https://githubcom/jts/nanopolish [Accessed January 10, 2019]. 2018;

26. Chen S, Zhou Y, Chen Y, Gu J. fastp: an ultra-fast all-in-one FASTQ preprocessor [Internet]. Vol. 34, Bioinformatics. 2018. p. i884–90. Available from: http://dx.doi.org/10.1093/bioinformatics/bty560

27. Bushnell B. BBMap. BBMap short read aligner, and other bioinformatic tools. 2014;

28. Hadfield J, Megill C, Bell SM, Huddleston J, Potter B, Callender C, et al. Nextstrain: real-time tracking of pathogen evolution. Bioinformatics. 2018 Dec 1;34(23):4121–3.

29. Rambaut A, Holmes EC, Hill V, O’Toole Á, McCrone JT, Ruis C, et al. A dynamic nomenclature proposal for SARS-CoV-2 to assist genomic epidemiology [Internet]. Available from: http://dx.doi.org/10.1101/2020.04.17.046086

30. O’Toole A, McCrone JT. Phylogenetic Assignment of Named Global Outbreak LINeages [Internet]. 2020 [cited 2020 Jun 18]. Available from: https://github.com/hCoV-2019/pangolin

31. Katoh K. MAFFT: a novel method for rapid multiple sequence alignment based on fast Fourier transform [Internet]. Vol. 30, Nucleic Acids Research. 2002. p. 3059–66. Available from: http://dx.doi.org/10.1093/nar/gkf436

32. De Maio N, Walker C, Borges R, Weilguny L, Slodkowicz G, Goldman N. Issues with SARS-CoV-2 sequencing data [Internet]. 2020 [cited 2020 Jun 16]. Available from: https://virological.org/t/issues-with-sars-cov-2-sequencing-data/473

33. Hasegawa M, Kishino H, Yano T-A. Dating of the human-ape splitting by a molecular clock of mitochondrial DNA [Internet]. Vol. 22, Journal of Molecular Evolution. 1985. p. 160–74. Available from: http://dx.doi.org/10.1007/bf02101694

34. Minh BQ, Schmidt HA, Chernomor O, Schrempf D, Woodhams MD, von Haeseler A, et al. IQ-TREE 2: New Models and Efficient Methods for Phylogenetic Inference in the Genomic Era. Mol Biol Evol. 2020 May 1;37(5):1530–4.

35. Guindon S, Dufayard J-F, Lefort V, Anisimova M, Hordijk W, Gascuel O. New algorithms and methods to estimate maximum-likelihood phylogenies: assessing the performance of PhyML 3.0. Syst Biol. 2010 May;59(3):307–21.

36. Sagulenko P, Puller V, Neher RA. TreeTime: Maximum-likelihood phylodynamic analysis. Virus Evol. 2018 Jan;4(1):vex042.

37. To T-H, Jung M, Lycett S, Gascuel O. Fast Dating Using Least-Squares Criteria and Algorithms. Syst Biol. 2016 Jan;65(1):82–97.

38. Organization WH, Organization WH, Others. Coronavirus disease (COVID-2019) situation reports. 2020.

39. World Population Prospects - Population Division - United Nations [Internet]. [cited 2020 Jun 19]. Available from: https://population.un.org/wpp/Download/Metadata/Documentation/

40. Candido DDS, Watts A, Abade L, Kraemer MUG, Pybus OG, Croda J, et al. Routes for COVID-19 importation in Brazil. J Travel Med [Internet]. 2020 May 18;27(3). Available from: http://dx.doi.org/10.1093/jtm/taaa042

41. Comunicado sobre vuelos de carácter humanitario del 2 al 12 de julio [Internet]. Cancillería. 2020 [cited 2020 Jun 19]. Available from: https://www.cancilleria.gov.co/newsroom/publiques/comunicado-vuelos-caracter-humanitario-2-12-julio

42. Sun K, Chen J, Viboud C. Early epidemiological analysis of the coronavirus disease 2019 outbreak based on crowdsourced data: a population-level observational study [Internet]. Vol. 2, The Lancet Digital Health. 2020. p. e201–8. Available from: http://dx.doi.org/10.1016/s2589-7500(20)30026-1

43. Castillo AE, Parra B, Tapia P, Acevedo A, Lagos J, Andrade W, et al. Phylogenetic analysis of the first four SARS-CoV-2 cases in Chile. J Med Virol [Internet]. 2020 Mar 29; Available from: http://dx.doi.org/10.1002/jmv.25797

44. da Silva Candido D, Claro IM, de Jesus JG, de Souza WM, Moreira FRR, Dellicour S, et al. Evolution and epidemic spread of SARS-CoV-2 in Brazil. medRxiv. 2020 Jun 12;2020.06.11.20128249.

45. Candido DS, Claro IM, de Jesus JG, Souza WM, Moreira FRR, Dellicour S, et al. Evolution and epidemic spread of SARS-CoV-2 in Brazil. Science [Internet]. 2020 Jul 23; Available from: http://dx.doi.org/10.1126/science.abd2161

46. Zhang L, Jackson CB, Mou H, Ojha A, Rangarajan ES, Izard T, et al. The D614G mutation in the SARS-CoV-2 spike protein reduces S1 shedding and increases infectivity. bioRxiv [Internet]. 2020 Jun 12; Available from: http://dx.doi.org/10.1101/2020.06.12.148726

47. Korber B, Fischer WM, Gnanakaran S, Yoon H, Theiler J, Abfalterer W, et al. Tracking Changes in SARS-CoV-2 Spike: Evidence that D614G Increases Infectivity of the COVID-19 Virus. Cell [Internet]. 2020 Jul 3; Available from: http://dx.doi.org/10.1016/j.cell.2020.06.043

48. Volz EM, Hill V, McCrone JT, Price A, Jorgensen D, O’Toole A, et al. Evaluating the effects of SARS-CoV-2 Spike mutation D614G on transmissibility and pathogenicity. medRxiv. 2020 Aug 4;2020.07.31.20166082.

49. Salazar C, Díaz-Viraqué F, Pereira-Gómez M, Ferrés I, Moreno P, Moratorio G, et al. Multiple introductions, regional spread and local differentiation during the first week of COVID-19 epidemic in Montevideo, Uruguay [Internet]. 2020 [cited 2020 Aug 10]. p. 2020.05.09.086223. Available from: https://www.biorxiv.org/content/10.1101/2020.05.09.086223v1

50. da Silva Candido D, Watts A, Abade L, Kraemer MUG, Pybus OG, Croda J, et al. Routes for COVID-19 importation in Brazil. medRxiv. 2020;

